# Age-specific effects of body size on fracture risk in later life: A lifecourse Mendelian randomization study

**DOI:** 10.1101/2021.12.06.21267379

**Authors:** Grace M. Power, Jon H. Tobias, Timothy M. Frayling, Jess Tyrrell, April Hartley, Jon Heron, George Davey Smith, Tom G. Richardson

**Affiliations:** MRC Integrative Epidemiology Unit, Population Health Sciences, Bristol Medical School, University of Bristol, Bristol, UK; Musculoskeletal Research Unit, Translational Health Sciences, Bristol Medical School, University of Bristol, Bristol, UK; Genetics of Complex Traits, College of Medicine and Health, University of Exeter, Exeter, UK; NIHR Bristol Biomedical Research Centre Bristol, University Hospitals Bristol and Weston NHS Foundation Trust, University of Bristol, Bristol, UK; Department of Genetics, Novo Nordisk Research Centre Oxford, Oxford, UK

## Abstract

Musculoskeletal conditions, including fractures, can have severe and long-lasting consequences. Higher body mass index in adulthood is widely acknowledged to be protective for most fracture sites, indicated through previous clinical and epidemiological observational research. However, the association between weight and bone health is complex and sources of bias, induced by confounding factors, may have distorted earlier findings. Employing a lifecourse Mendelian randomization (MR) approach by using genetic instruments to separate effects at different life stages, this investigation aims to explore how prepubertal and adult body size independently influence fracture risk in later life.

Using data from a large UK-based prospective cohort, univariable and multivariable MR with inverse variance weighted meta-analysis were conducted to simultaneously estimate the effects of age-specific genetic proxies for body size (n=453,169) on the odds of fracture in later life (n=416,795). A two-step MR framework was additionally applied to elucidate potential mediators. Univariable and multivariable MR indicated strong evidence that higher body size in childhood reduced fracture risk in later life (OR, 95% CI: 0.89, 0.82 to 0.96, P=0.005 and OR, 95% CI: 0.76, 0.69 to 0.85, P=1×10^−6^, respectively). Conversely, higher body size in adulthood increased fracture risk (OR, 95% CI: 1.08, 1.01 to 1.16, P=0.023 and OR, 95% CI: 1.26, 1.14 to 1.38, P=2×10^−6^, respectively). Two-step MR analyses suggested that the effect of higher body size in childhood on reduced fracture risk was mediated by its influence on higher estimated bone mineral density (eBMD) in adulthood.

This investigation provides novel evidence that higher body size in childhood has a direct effect on reduced fracture risk in later life through its influence on increased eBMD. Results indicate that higher body size in adulthood is a risk factor for fractures, opposing findings from earlier research. Protective effect estimates previously observed are likely attributed to childhood effects.

## Introduction

Musculoskeletal conditions are a leading cause of disability worldwide, affecting nearly 2 billion people (1). Associated injuries, including fractures and falls, can lead to serious and long-lasting effects, particularly in later life (2). With hip fracture predicted to incur an annual worldwide cost of US$132 billion by 2050 (3), not only the social, but economic burden of these health states make prevention of such conditions an important public health goal (4).

Considerable changes occur in body composition over the lifecourse (5). Bone mass is accrued until peak bone mass is reached in the third decade of life (6). It then remains stable until menopause in women, and later life in men, where sex steroid deficiency begins to drive cortical bone loss (7, 8). Lean mass additionally increases during growth in childhood, remaining largely stable following puberty until falling later in life (9). Conversely, body fat tends to rise in older age groups, with obesity prevalence peaking in individuals aged 60 to 69 years in high-income countries (10, 11). Several meta-analyses have pointed to a complex association between body mass index (BMI) and fracture risk (12-14). Although higher BMI in adulthood is widely acknowledged to be a protective factor for most sites of fragility fracture (12, 13), studies have shown conflicting results with some evidence suggesting that obesity may be related to an increased risk of fracture (14, 15). Mendelian randomization (MR) investigations exploit the quasi-random assortment of genetic variants independent of other traits to mitigate against false inferences resulting from confounding and reverse causality (16-18). Using the principles of MR, investigations have indicated that higher adiposity increases bone mineral density (BMD) in childhood (16) and that low BMD increases the risk of fracture (19). Moreover, a population-based birth cohort study supported by a subsequent MR investigation identified fat mass as a positive determinant of bone mass and size in prepubertal children (20, 21), pointing to the positive effects of loading on bone formation at a young age (16). Fat mass may stimulate bone growth in childhood through a direct mechanical action of increased load (22), or indirectly by association with increased lean mass, since both are strongly correlated across the whole range of body mass (23). It is therefore plausible that the protective effect estimates observed between BMI in adults and fracture risk in later life, could be attributed to the effects of higher body size that may have exerted an influence on the skeleton in childhood.

Separating the effects of body size at different stages of the lifecourse is challenging, particularly due to the influence of confounding factors, often afflicting observational studies. This is a key motivation behind using a lifecourse MR approach, which intends to estimate the causal effect of time-varying modifiable risk factors under specific assumptions; the instrumental variables used must i) associate with the exposure of interest conditional on the other exposures (the ‘relevance’ assumption), ii) not affect the outcome except through the exposures (the ‘exclusion restriction’ assumption) and iii) be independent of all confounders, both observed and unobserved, of any of the exposures and outcome (the ‘exchangeability’ assumption) (24). The aim of this investigation was to apply this approach to explore how body size at different stages in the lifecourse modifies the risk of fractures in later life. First, we conducted univariable MR to estimate the ‘total’ effect of early life body size on the odds of fracture (**Figure 1A**). Multivariable MR, an extension of MR that employs multiple genetic variants associated with multiple measured risk factors to simultaneously estimate the causal effect of each exposure on the outcome of interest, was then employed (25). This was to estimate the ‘direct’ and ‘indirect’ effects of childhood body size on the odds of fracture in later life, by taking adulthood body size into account **(Figure 1B**). We then applied the principles of MR in a two-step framework (26) to assess potential mediators; by examining calcium and vitamin D levels, as well as BMD in adulthood, estimated by quantitative ultrasound of the heel calcaneus (hereafter, “eBMD”) (**Figure 1C**). Given the literature, suggesting that effects of weight are often sex-specific (27), we explored these questions separately by sex. We examined three hormones (bioavailable testosterone, total testosterone, and sex hormone binding globulin (SHBG)) shown to be related to BMI and bone health, as potential mediators between childhood BMI and both fracture risk in later life and eBMD, in the sex-stratified population (28-31).

**Figure 1.**
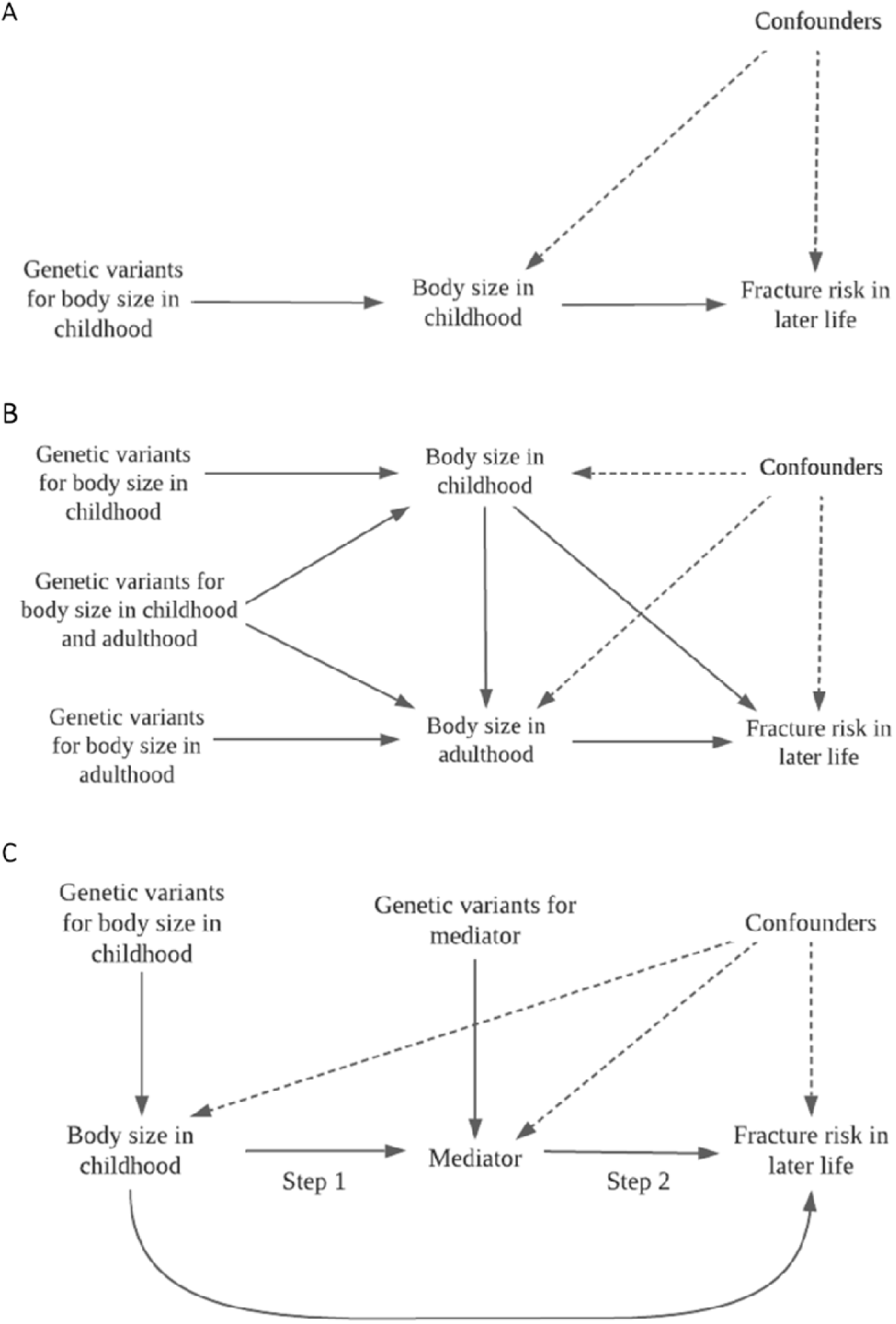
Directed acyclic graphs indicating three scenarios to explain the causal effect between childhood body size and fracture risk in later life. (A) Univariable Mendelian randomization measuring the total effect of body size in childhood on the odds of fracture in later life. (B) Multivariable Mendelian randomization measuring the direct and indirect effect of body size in childhood and adulthood on the odds of fracture in later life. (C) Two-step Mendelian randomization measuring the total effect of body size in childhood on the mediator (Step 1) and the total effect of the mediator on the odds of fracture in later life (Step 2), allowing the measurement of th indirect effect of body size in childhood on the odds of fracture in later life via the mediator, *e*.*g*., estimated bone mineral density.

## Materials and methods

### Data resources

Genetic variants strongly associated with childhood and adult body size (using P < 5×10^−8^ and r^2^ < 0.001) were identified in a large-scale genome-wide association study (GWAS), previously undertaken in the UK Biobank study on 453,169 individuals, adjusting for age, sex, and genotyping chip (32, 33). UK Biobank data were collected between 2006 and 2010 on individuals aged between 40 and 69 years old at baseline, from clinical examinations, assays of biological samples, detailed information on self-reported health characteristics, and genome-wide genotyping, using a prospective cohort study design (33). The childhood body size measure applied in this study, utilised recall questionnaire data, involving responses from adult participants who were asked whether, compared to the average, they were ‘thinner’, ‘about average’ or ‘plumper’, when they were aged 10 years old. The adult body size variable was derived using clinically measured body mass index (BMI) data (mean age 56.5 years). It was then separated into a 3-tier variable using the same categories as the childhood body size measure; “thinner” (21.1kg/m^2^-25kg/m^2^), “about average” (25kg/m^2^-31.7kg/m^2^) and “plumper” (31.7kg/m^2^-59.9kg/m^2^) (34, 35). Individuals that did not have data for both childhood and adult body size were excluded from analyses and a genetic correlation coefficient of rG=0.61 was previously calculated between these two measures (36). In addition, these scores have been independently validated in two distinct cohorts, providing verification that these genetic instruments can reliably separate childhood and adult body size. Furthermore, comparing the genetic correlation between the childhood body size GWAS with a recent GWAS of measured childhood BMI, provided strong evidence of validation using LD score regression (rg=0.96) (37).

A previously published GWAS on individuals in the UK Biobank for the main outcome, fractures in later life, was used (n=416,795) (38). This excluded fractures of the skull, face, hands and feet, pathological fractures due to malignancy, atypical femoral fractures, periprosthetic, and healed fracture codes and a full list of ICD10 codes used have been reported previously (38). Effect estimates derived from results indicate an additive change in the odds of each change in weight category in childhood and adult body size (36). To generate genetic instruments for birthweight (n=261,932), GWAS were undertaken on UK Biobank individuals with adjustment for age, sex and genotyping chip. We used a linear mixed model to account for genetic relatedness and geographical structure in UK Biobank as undertaken with the BOLT-LMM software. Birthweight was kept as a continuous trait given that it was not available in the full sample and rank-based inverse normal transformed, to ensure values lay within accepted limits assuming a normal distribution. We additionally ran GWAS for several potential mediators using UK Biobank data with the application of the same analysis pipeline stipulated above. Mediators included genetic predisposition to increased serum calcium in nmol/L (n=432,151), vitamin D level in nmol/L (n=449,913) and eBMD, derived from heel ultrasound (n=278,932), in the total population, as well as bioavailable testosterone (female n=206,604, male n=207,470), total testosterone (female n=180,386, male n=184,025), and SHBG (female n=222,491, male n=205,646), in the sex-stratified population. We standardised the distribution of these variables to have a mean of 0 and standard deviation of 1 and results are quantified as standard deviation change. In addition, in sensitivity analyses this study estimated effects on sex (n=361,194) to assess participation bias, as well as total body fat mass (n=453,957) and total body fat-free mass (n=454,669) which were made into indices by dividing by height (m)^2^ using UK Biobank data (S1 Table).

Conducting MR using overlapping sets of participant samples has been shown to bias in the direction of the observational association between the risk factor and outcome (39). We therefore used previously developed formulae implemented in a web application (https://sb452.shinyapps.io/overlap/) to calculate expected bias and Type 1 error rate under the null for genetically proxied childhood body size and odds of fracture in later life (40). Data used to generate output is in S2 Table. Estimated bias due to sample overlap is presented in S3 Table.

The UK Biobank study have obtained ethics approval from the Research Ethics Committee (REC; approval number: 11/NW/0382) and informed consent from all participants enrolled in UK Biobank. Estimates were derived using data from the UK Biobank (app #76538).

### Statistical analysis

Univariable MR was initially conducted to estimate the ‘total’ effects of genetically predicted childhood body size and adult body size on fractures in later life. Firstly, the inverse variance weighted method (IVW) was employed, which takes SNP-outcome estimates and regresses them on the SNP-exposure associations (41). Complementary methods, namely weighted median and MR-Egger were subsequently used to assess the robustness of the univariable results obtained. This provided more robust to MR assumptions compared to the IVW method, including horizontal pleiotropy, whereby genetic variants influence multiple traits or disease outcomes via independent biological pathways (42). Multivariable IVW MR was used to calculate the direct and indirect effects of childhood body size and adult body size, simultaneously, on fractures in later life, accounting for either adult body size or childhood body size, respectively i.e., the exposure variables that were not considered the main exposure of interest, in each model (24). Genetic estimates for our exposures were harmonized with mediators and the disease outcome using the ‘TwoSampleMR’ R package. Forest plots in this paper were generated using the R package ‘ggplot2’ (43). These analyses were undertaken using R (version 3.5.1).

Birthweight was investigated as a third exposure related to body size in i) univariable MR, assessing the ‘total’ effect of birthweight on the outcome, fractures in later life, ii) multivariable MR assessing the ‘direct’ and ‘indirect’ effect of birthweight on the outcome, by taking childhood and adult body size into account, and iii) in both childhood and adult body size multivariable MR models, accounting for birthweight, to determine whether any effects of childhood and adult body size observed are a result of weight in very early life (44). A combination of foetal- and maternal-specific mechanisms and tissues have been identified in the regulation of birthweight, with some mechanisms involving directionally opposing effects in the foetus and mother (45). Therefore, this investigation of birthweight was not to determine the effects of parental factors on fracture risk in later life, but to exclude the possibility that very early life body size is an explanation of the childhood BMI effect.

#### Two-step Mendelian Randomization

To investigate the possible mechanisms by which body size affects fractures and determine potential intermediate traits, two-step MR was conducted (26). This was achieved by, i) assessing the separate effects of genetically predicted childhood body size and adult body size on each of the potential non-sex-specific mediators; eBMD, calcium, and vitamin D, as outcomes, and then, ii) assessing each of these mediators as exposures on the outcome, fractures in later life. Multivariable IVW MR analyses were additionally computed to calculate the direct and indirect effects of childhood body size and adult body size, simultaneously, on mediators of interest.

#### Sex-stratification

Univariable and multivariable MR analyses were conducted in sex-stratified groups; female or male, assigned at birth. Two-sample MR was subsequently computed to investigate bioavailable testosterone, total testosterone, and SHBG, to determine any potential hormonal sex-specific mechanisms of action between body size and eBMD as well as fractures in later life.

### Sensitivity analyses

#### Body composition measures

To establish whether results indicate a true causal effect of childhood body size, that does not discriminate between adiposity and lean mass, on fracture risk in later life, we explored the effects of childhood and adult body size on fat mass index and fat-free mass index in a univariable and multivariable MR setting. In addition, we conducted multivariable MR analyses to assess the relationship between childhood body size and fracture risk in later life, accounting for fat mass index and fat-free mass index to estimate potential mediation. This was also to maintain an estimate with suitable comparability to the adult measure used (a genetic proxy for BMI).

#### Sex-differential participation bias

We investigate the potential for artefactual associations as a result of sex-differential participation, whereby childhood or adult body size may have led males and females to differentially participate in the UK Biobank study. This was achieved through estimating variants associated with the traits, childhood and adult body size, and sex (46).

#### Family-based analyses

Findings from MR analyses of unrelated individuals may be biased as a result of uncontrolled confounding from familial effects: dynastic effects, assortative mating, or population stratification (47). It has been argued that within-family genetic association estimates, for example, those acquired from samples of siblings, may allow more accurate estimates of direct genetic effects since these are unaffected by demography and indirect genetic effects of parents (48). Population (between-family) and within-sibship (within-family) estimates (n=39,507) were therefore generated to ensure the direction of our estimates were not a result of dynastic effects, assortative mating, or population stratification.

## Results

Univariable analyses indicated evidence that higher genetically predicted childhood body size reduced the odds of fractures in later life (IVW OR, 95% CI: 0.89, 0.82 to 0.96, P=0.005). Multivariable MR analyses showed strong evidence of an effect between higher genetically predicted childhood body size and fractures in later life, after accounting for adult body size (OR, 95% CI: 0.76, 0.69 to 0.85, P=1×10^−6^). There was some evidence of an association between higher genetically predicted adult body size and the odds of fractures in later life (OR, 95% CI: 1.08, 1.01 to 1.16, P=0.023). There was additionally strong evidence of an effect between higher genetically predicted adult body size and fractures in later life, after accounting for childhood body size (OR, 95% CI: 1.26, 1.14 to 1.38, P=2×10^−6^) (Table 1, Figure 2). Consistent patterns of association were apparent when using the weighted median method employed for robustness in univariable analyses. Results using the MR-Egger method did not provide evidence that horizontal pleiotropy was driving these effects in childhood, with effect estimates similar when using the IVW method, however, those for adult body size demonstrated the reverse direction of effect, suggesting the effect of adult body size may be prone to horizontal pleiotropy (Table 1).

**Table 1.**
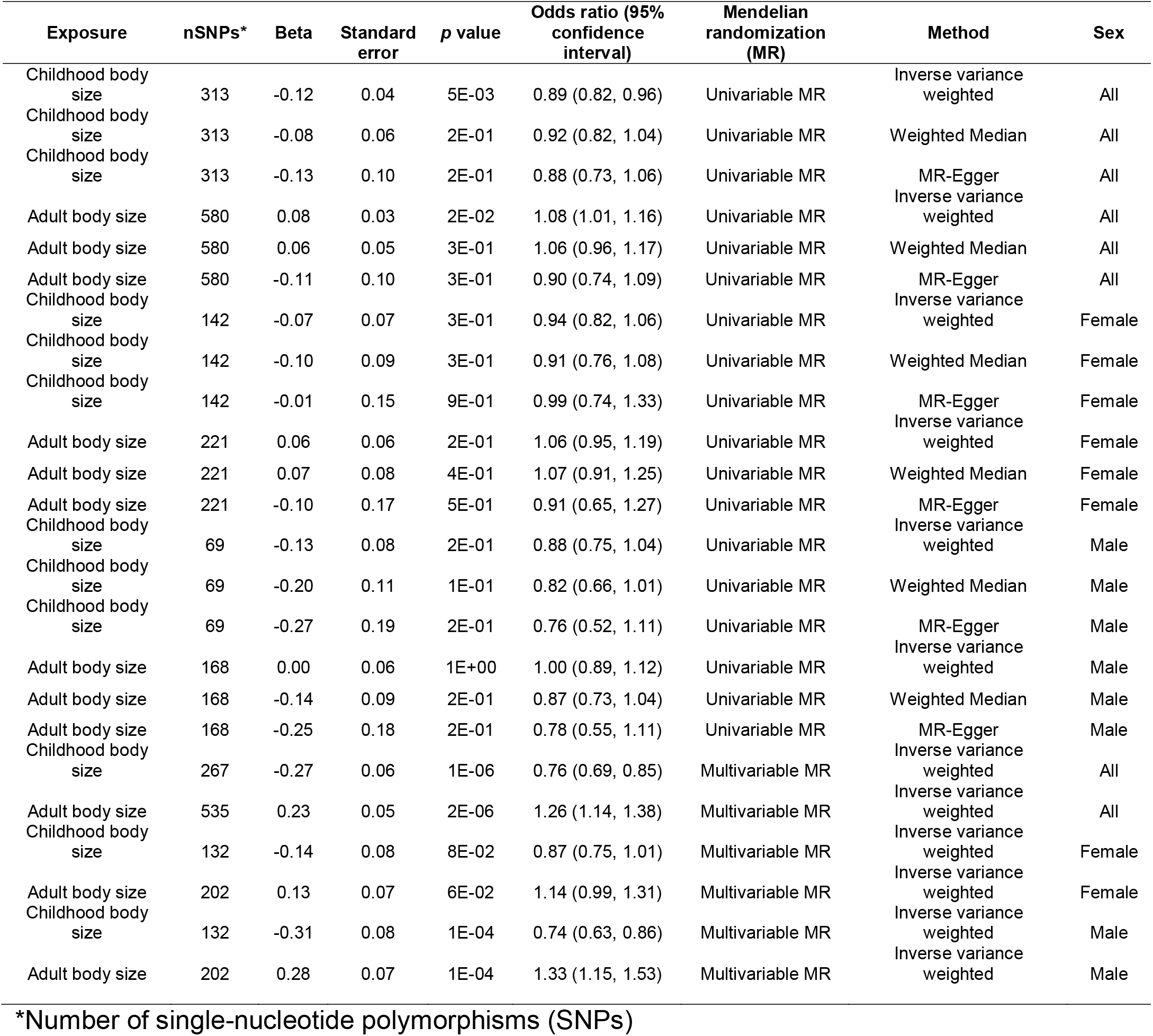
Univariable and multivariable Mendelian randomization analyses for childhood and adult body size onto fracture risk in later life

**Figure 2.**
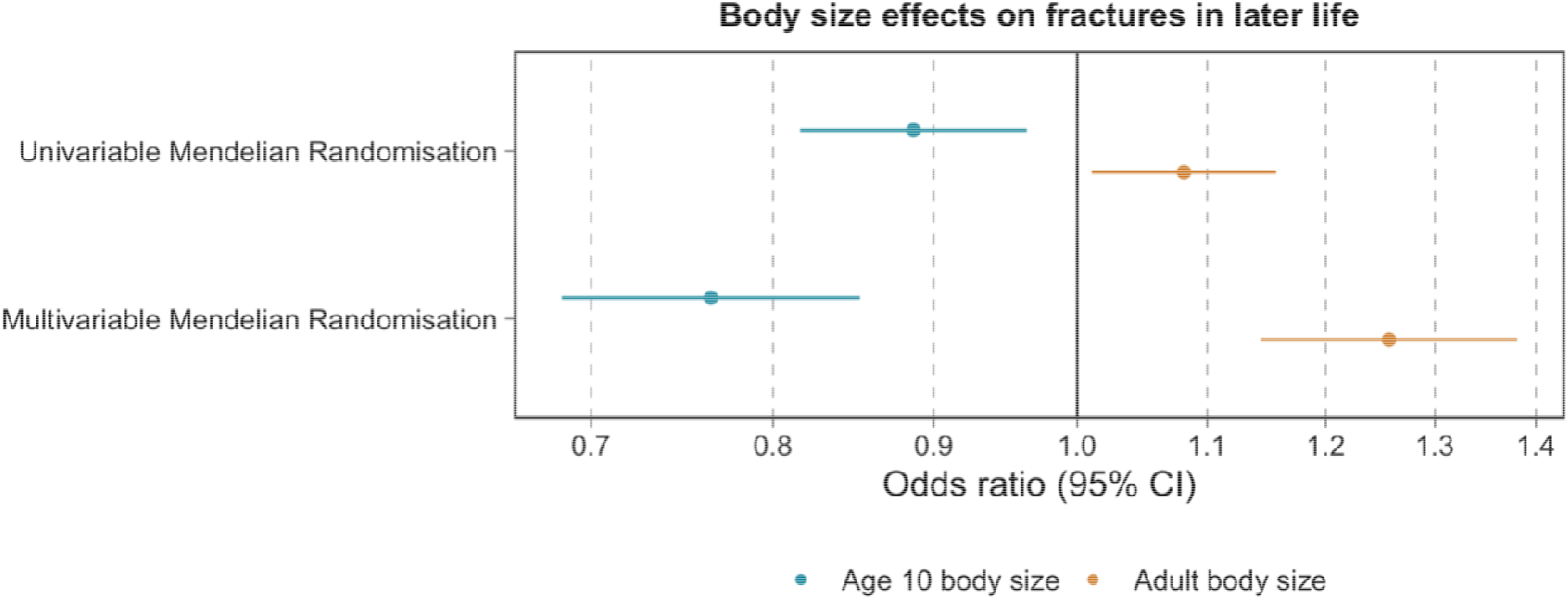
Univariable and multivariable Mendelian randomization for childhood and adult body size onto the outcome measure, fractures in later life. Dots filled-in indicate some to very strong statistical evidence of an association.

In univariable analyses, there was some evidence of an effect between genetically predicted higher birthweight and increase in fractures in later life (OR, 95% CI: 1.08, 1.01 to 1.16, P=0.036). In multivariable analyses, there was evidence of an effect between genetically predicted higher birthweight and increase in fractures in later life (OR, 95% CI: 1.03, 1.01 to 1.17, P=0.003), accounting for childhood and adult body size. There was strong evidence of an effect between childhood body size and fractures in later life, after accounting for adult body size and birthweight (OR, 95% CI: 0.78, 0.69 to 0.88, P=7×10^−5^). There was additionally strong evidence of an effect between adult body size and fractures in later life (OR, 95% CI: 1.21, 1.09 to 1.35, P=3×10^−4^), after accounting for childhood body size and birthweight (Table 2).

**Table 2.** Univariable and multivariable Mendelian randomization analyses for birthweight, childhood and adult body size onto fracture risk in later life and estimated bone mineral density

| Exposure | Outcome | nSNPs* | Beta | Standard error | p value | Odds ratio (95% confidence interval) | Mendelian randomization (MR) |
| --- | --- | --- | --- | --- | --- | --- | --- |
| Birthweight | Fracture risk in later life | 174 | 0.08 | 0.04 | 3E-02 | 1.08 (1.01, 1.16) | Univariable MR |
| Childhood body size | Fracture risk in later life | 313 | -0.12 | 0.04 | 5E-03 | 0.89 (0.82, 0.96) | Univariable MR |
| Adult body size | Fracture risk in later life | 580 | 0.08 | 0.03 | 2E-02 | 1.08 (1.01, 1.16) | Univariable MR |
| Birthweight | eBMD | 174 | -0.08 | 0.03 | 1E-02 | (-0.14, 0.02) | Univariable MR |
| Childhood body size | eBMD | 313 | 0.26 | 0.03 | 3E-16 | (0.20, 0.33) | Univariable MR |
| Adult body size | eBMD | 580 | 0.20 | 0.02 | 1E-17 | (0.15, 0.25) | Univariable MR |
| Birthweight | Fracture risk in later life | 148 | 0.10 | 0.03 | 3E-03 | 1.10 (1.03, 1.17) | Multivariable MR |
| Childhood body size | Fracture risk in later life | 203 | -0.25 | 0.06 | 7E-05 | 0.78 (0.69, 0.88) | Multivariable MR |
| Adult body size | Fracture risk in later life | 396 | 0.19 | 0.05 | 3E-04 | 1.21 (1.09, 1.35) | Multivariable MR |
| Birthweight | eBMD | 148 | -0.11 | 0.02 | 1E-05 | (-0.16, -0.06) | Multivariable MR |
| Childhood body size | eBMD | 203 | 0.21 | 0.05 | 2E-05 | (0.11, 0.30) | Multivariable MR |
| Adult body size | eBMD | 396 | 0.11 | 0.04 | 1E-02 | (0.02, 0.19) | Multivariable MR |

\*Number of single-nucleotide polymorphisms (SNPs)

### Two-step Mendelian randomization

Among the mediators assessed in the total population, strong evidence of an effect was observed in both MR steps for childhood and adult body size and eBMD. An increase per one standard deviation in genetically predicted childhood and adult body size indicated an increase in eBMD (beta, 95% CI: 0.26, 0.20 to 0.33, P=3×10^−16^ and 0.20, 0.16 to 0.25, P=1×10^−17^, respectively). In multivariable MR, the effect estimate slightly weakened when estimating childhood body size on eBMD (beta, 95% CI: 0.20, 0.12 to 0.27, P=3×10^−7^) and substantially when estimating adult body size on eBMD (beta, 95% CI: 0.09, 0.02 to 0.15, P=0.008) (Figure 3). In addition, an increase in eBMD demonstrated a decrease in fractures in later life (OR, 95% CI: 0.62, 0.60 to 0.64, P=5×10^−150^) (Table 3).

**Figure 3.**
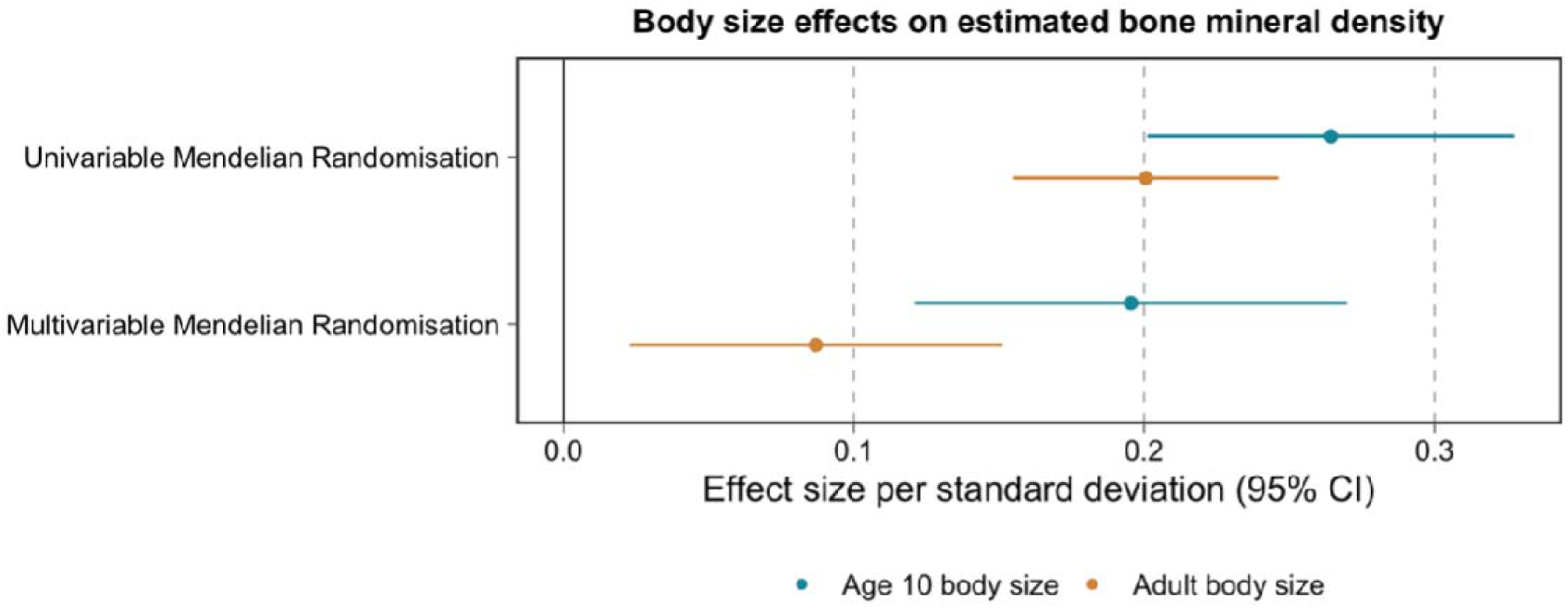
Univariable and multivariable Mendelian randomization for childhood and adult body size onto estimated bone mineral density. Dots filled-in indicate some to very strong statistical evidence of an association.

**Table 3.** Two-step Mendelian randomization analyses for childhood and adult body size onto potential mediators and potential mediators onto fracture risk in later life and estimated bone mineral density

| Exposure | Mediator/<br>Outcome | nSNPs* | Beta | Standard<br>error | p value | Odds ratio (95%<br>confidence<br>interval) | Mendelian<br>randomization<br>(MR) | Method | Sex |
| --- | --- | --- | --- | --- | --- | --- | --- | --- | --- |
| Childhood body size | eBMD | 313 | 0.26 | 0.03 | 3E-16 | (0.20, 0.33) | Univariable MR | Inverse variance weighted | All |
| Childhood body size | eBMD | 313 | 0.25 | 0.03 | 7E-21 | (0.20, 0.30) | Univariable MR | Weighted median | All |
| Childhood body size | eBMD | 313 | 0.36 | 0.07 | 1E-06 | (0.21, 0.50) | Univariable MR | MR Egger | All |
| Childhood body size | eBMD | 142 | 0.18 | 0.04 | 7E-06 | (0.10, 0.26) | Univariable MR | Inverse variance weighted | Female |
| Childhood body size | eBMD | 142 | 0.22 | 0.04 | 5E-10 | (0.15, 0.29) | Univariable MR | Weighted median | Female |
| Childhood body size | eBMD | 142 | 0.27 | 0.09 | 4E-03 | (0.09, 0.44) | Univariable MR | MR Egger | Female |
| Childhood body size | eBMD | 69 | 0.36 | 0.06 | 3E-08 | (0.23, 0.49) | Univariable MR | Inverse variance weighted | Male |
| Childhood body size | eBMD | 69 | 0.37 | 0.05 | 6E-13 | (0.27, 0.47) | Univariable MR | Weighted median | Male |
| Childhood body size | eBMD | 69 | 0.44 | 0.15 | 4E-03 | (0.15, 0.73) | Univariable MR | MR Egger | Male |
| Adult body size | eBMD | 580 | 0.20 | 0.02 | 1E-17 | (0.15, 0.25) | Univariable MR | Inverse variance weighted | All |
| Adult body size | eBMD | 580 | 0.22 | 0.02 | 1E-29 | (0.18, 0.26) | Univariable MR | Weighted median | All |
| Adult body size | eBMD | 580 | 0.24 | 0.07 | 3E-04 | (0.11, 0.37) | Univariable MR | MR Egger | All |
| Adult body size | eBMD | 221 | 0.16 | 0.04 | 9E-06 | (0.09, 0.24) | Univariable MR | Inverse variance weighted | Female |
| Adult body size | eBMD | 221 | 0.16 | 0.03 | 3E-07 | (0.10, 0.23) | Univariable MR | Weighted median | Female |
| Adult body size | eBMD | 221 | 0.22 | 0.11 | 5E-02 | (0.00, 0.44) | Univariable MR | MR Egger | Female |
| Adult body size | eBMD | 168 | 0.34 | 0.04 | 4E-14 | (0.25, 0.42) | Univariable MR | Inverse variance weighted | Male |
| Adult body size | eBMD | 168 | 0.37 | 0.05 | 7E-16 | (0.28, 0.45) | Univariable MR | Weighted median | Male |
| Adult body size | eBMD | 168 | 0.52 | 0.13 | 2E-04 | (0.26, 0.78) | Univariable MR | MR Egger | Male |
| Childhood body size | Calcium | 313 | -0.10 | 0.02 | 6E-07 | (-0.15, -0.06) | Univariable MR | Inverse variance weighted | All |
| Childhood body size | Calcium | 313 | -0.09 | 0.02 | 1E-05 | (-0.13, -0.05) | Univariable MR | Weighted median | All |
| Childhood body size | Calcium | 313 | -0.11 | 0.05 | 2E-02 | (-0.20, -0.02) | Univariable MR | MR Egger | All |
| Childhood body size | Vitamin D | 313 | -0.07 | 0.02 | 5E-05 | (-0.10, -0.04) | Univariable MR | Inverse variance weighted | All |
| Childhood body size | Vitamin D | 313 | -0.06 | 0.02 | 2E-03 | (-0.10, -0.02) | Univariable MR | Weighted median | All |
| Childhood body size | Vitamin D | 313 | -0.11 | 0.04 | 3E-03 | (-0.19, -0.04) | Univariable MR | MR Egger | All |
| Adult body size | Calcium | 580 | -0.05 | 0.02 | 6E-03 | (-0.08, -0.01) | Univariable MR | Inverse variance weighted | All |
| Adult body size | Calcium | 580 | -0.07 | 0.02 | 6E-05 | (-0.10, -0.03) | Univariable MR | Weighted median | All |
| Adult body size | Calcium | 580 | -0.09 | 0.05 | 7E-02 | (-0.20, 0.01) | Univariable MR | MR Egger | All |
| Adult body size | Vitamin D | 580 | -0.18 | 0.01 | 1E-37 | (-0.21, -0.15) | Univariable MR | Inverse variance weighted | All |
| Adult body size | Vitamin D | 580 | -0.19 | 0.02 | 3E-32 | (-0.22, -0.16) | Univariable MR | Weighted median | All |
| Adult body size | Vitamin D | 580 | -0.18 | 0.04 | 2E-05 | (-0.26, -0.10) | Univariable MR | MR Egger | All |
| Childhood body size | eBMD | 267 | 0.20 | 0.04 | 3E-07 | (0.12, 0.27) | Multivariable MR | Inverse variance weighted | All |
| Adult body size | eBMD | 535 | 0.09 | 0.03 | 8E-03 | (0.02, 0.15) | Multivariable MR | Inverse variance weighted | All |
| Childhood body size | eBMD | 132 | 0.21 | 0.04 | 5E-07 | (0.13, 0.29) | Multivariable MR | Inverse variance weighted | Female |
| Adult body size | eBMD | 202 | 0.04 | 0.04 | 3E-01 | (-0.03, 0.12) | Multivariable MR | Inverse variance weighted | Female |
| Childhood body size | eBMD | 58 | 0.20 | 0.07 | 8E-03 | (0.05, 0.35) | Multivariable MR | Inverse variance weighted | Male |
| Adult body size | eBMD | 161 | 0.14 | 0.06 | 3E-02 | (0.02, 0.27) | Multivariable MR | Inverse variance weighted | Male |
| Childhood body size | Calcium | 267 | -0.11 | 0.03 | 1E-04 | (-0.17, -0.05) | Multivariable MR | Inverse variance weighted | All |
| Adult body size | Calcium | 535 | 0.01 | 0.03 | 6E-01 | (-0.04, 0.06) | Multivariable MR | Inverse variance weighted | All |
| Childhood body size | Vitamin D | 267 | 0.07 | 0.02 | 3E-03 | (0.02, 0.11) | Multivariable MR | Inverse variance weighted | All |
| Adult body size | Vitamin D | 535 | -0.22 | 0.02 | 4E-30 | (-0.26, -0.18) | Multivariable MR | Inverse variance weighted | All |
| eBMD | Fractures | 548 | -0.48 | 0.02 | 5E-150 | 0.62 (0.60, 0.64) | Univariable MR | Inverse variance weighted | All |
| eBMD | Fractures | 548 | -0.48 | 0.02 | 3E-91 | 0.62 (0.59, 0.65) | Univariable MR | Weighted median | All |
| eBMD | Fractures | 548 | -0.54 | 0.04 | 9E-41 | 0.58 (0.54, 0.63) | Univariable MR | MR Egger | All |
| eBMD | Fractures | 323 | -0.51 | 0.02 | 2E-112 | 0.60 (0.57, 0.63) | Univariable MR | Inverse variance weighted | Female |
| eBMD | Fractures | 323 | -0.56 | 0.03 | 9E-77 | 0.57 (0.54, 0.61) | Univariable MR | Weighted median | Female |
| eBMD | Fractures | 323 | -0.59 | 0.05 | 1E-29 | 0.55 (0.51, 0.61) | Univariable MR | MR Egger | Female |
| eBMD | Fractures | 206 | -0.37 | 0.03 | 5E-40 | 0.69 (0.65, 0.73) | Univariable MR | Inverse variance weighted | Male |
| eBMD | Fractures | 206 | -0.47 | 0.04 | 1E-38 | 0.63 (0.58, 0.67) | Univariable MR | Weighted median | Male |
| eBMD | Fractures | 206 | -0.47 | 0.06 | 2E-13 | 0.62 (0.55, 0.70) | Univariable MR | MR Egger | Male |
| Calcium | Fractures | 355 | -0.01 | 0.02 | 8E-01 | 0.99 (0.95, 1.04) | Univariable MR | Inverse variance weighted | All |
| Calcium | Fractures | 355 | 0.01 | 0.03 | 7E-01 | 1.01 (0.95, 1.08) | Univariable MR | Weighted median | All |
| Calcium | Fractures | 355 | 0.01 | 0.05 | 8E-01 | 1.01 (0.92, 1.10) | Univariable MR | MR Egger | All |
| Vitamin D | Fractures | 131 | 0.02 | 0.03 | 5E-01 | 1.02 (0.96, 1.09) | Univariable MR | Inverse variance weighted | All |
| Vitamin D | Fractures | 131 | -0.02 | 0.05 | 8E-01 | 0.98 (0.89, 1.09) | Univariable MR | Weighted median | All |
| Vitamin D | Fractures | 131 | 0.00 | 0.05 | 1E+00 | 1.00 (0.91, 1.10) | Univariable MR | MR Egger | All |
| Calcium | eBMD | 355 | -0.04 | 0.02 | 4E-02 | (-0.07, 0.00) | Univariable MR | Inverse variance weighted | All |
| Calcium | eBMD | 355 | -0.04 | 0.02 | 2E-02 | (-0.06, -0.01) | Univariable MR | Weighted median | All |
| Calcium | eBMD | 355 | -0.02 | 0.03 | 5E-01 | (-0.09, 0.05) | Univariable MR | MR Egger | All |
| Vitamin D | eBMD | 131 | -0.01 | 0.02 | 6E-01 | (-0.06, 0.03) | Univariable MR | Inverse variance weighted | All |
| Vitamin D | eBMD | 131 | -0.04 | 0.02 | 1E-02 | (-0.07, -0.01) | Univariable MR | Weighted median | All |
| Vitamin D | eBMD | 131 | -0.02 | 0.03 | 5E-01 | (-0.09, 0.05) | Univariable MR | MR Egger | All |

There was strong evidence of an association between genetically predicted childhood body size and calcium (beta, 95% CI: -0.11, -0.15 to -0.06, P=6×10^−7^), and evidence of an effect between genetically predicted adult body size and calcium (beta, 95% CI: - 0.05, -0.09 to -0.01, P=0.006), however, little evidence of an association between calcium and fractures in later life (OR, 95% CI: 0.99, 0.95 to 1.04, P=0.813). Furthermore, there was strong evidence or an effect between genetically predicted childhood and adult body size on vitamin D (beta, 95% CI: -0.07, -0.10 to -0.04, P=5×10^−5^ and -0.18, -0.21 to -0.16, P=1×10^−37^) and little evidence of an effect between vitamin D and fractures in later life (OR, 95% CI: 1.02, 0.96 to 1.09, P=0.541) (Table 3, Figure 4).

**Figure 4.**
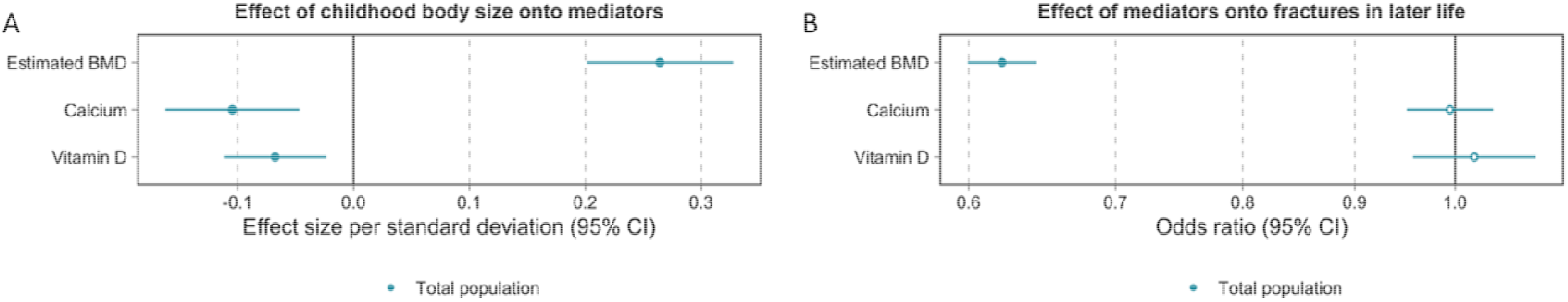
Two-step mendelian randomization results estimating (A) childhood body size on selected mediators and (B) selected mediators onto the outcome measure, fractures in later life, in the total population. Dots filled-in indicate some to very strong statistical evidence of an association.

### Sex-stratification

Upon stratification, there was strong evidence of an association between childhood body size and eBMD in females (beta, 95% CI: 0.18, 0.10 to 0.26, P=7×10^−6^) and in males (beta, 95% CI: 0.36, 0.23 to 0.49, P=3×10^−8^). After accounting for adult body size, there remained evidence of an increase in eBMD with central effect estimates more comparable between females and males (beta, 95% CI: 0.21, 0.13 to 0.29, P=5×10^−7^and beta, 95% CI: 0.20, 0.05 to 0.346, P=0.008, respectively). There was additionally strong evidence of an increase in eBMD and decrease in fractures in later life in females (OR, 95% CI: 0.60, 0.57 to 0.63, P=2×10^−112^) and males (OR, 95% CI: 0.69, 0.65 to 0.73, P=5×10^−40^) (Table 3). Consistent patterns of associations were observed using the weighted median method employed for robustness. In addition, results using the MR-Egger method did not provide evidence that horizontal pleiotropy was responsible for the estimates derived. Two-sample MR estimates computed to investigate bioavailable testosterone, total testosterone, and SHBG, as potential mechanisms of action between body size and fractures in later life are presented in Figure 5. Further estimates from two-sample analyses, including those which account for adult body size, are presented in S4 Table.

**Figure 5.**
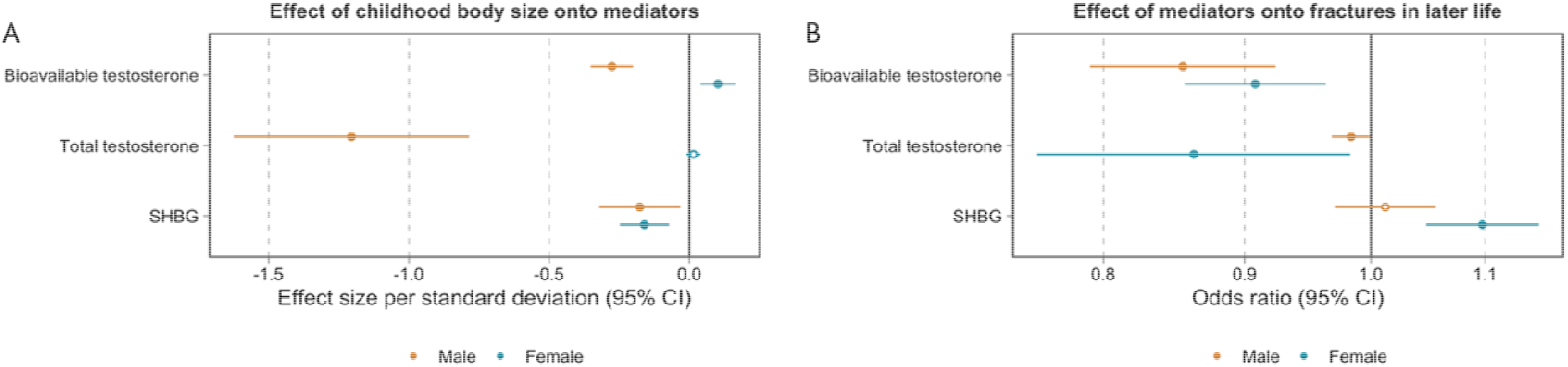
Two-step mendelian randomization results estimating (A) childhood body size on selected mediators and (B) selected mediators onto the outcome measure, fractures in later life, sex stratified. Dots filled-in indicate some to very strong statistical evidence of an association.

### Sensitivity analyses

#### Body composition measures

There was strong evidence of an effect between childhood and adult body size and fat-free mass index, measured in adulthood (beta, 95% CI: 0.75, 0.69 to 0.792 P=3×10^−195^ and 1.00, 0.98 to 1.02, P=1×10^−300^, respectively). Consistent patterns of associations were observed using the weighted median method employed for robustness and results using the MR-Egger method did not provide evidence that horizontal pleiotropy was responsible for the derived estimates (S5 Table). In multivariable MR analyses, whilst the beta reduced, there remained strong evidence of an association between genetically predicted childhood body size and fat-free mass index, after accounting for adult body size (beta, 95% CI: 0.18, 0.14 to 0.21, P=8×10^−23^). There was additionally strong evidence of an effect between higher genetically predicted adult body size and an increase in fat-free mass index, after accounting for childhood body size (beta, 95% CI: 0.91, 0.88 to 0.94, P=1×10^−300^). Furthermore, in univariable analyses, there was strong evidence of an effect between higher childhood and adult body size and increased fat mass index, measured in adulthood (beta, 95% CI: 0.78, 0.72 to 0.84, P=7×10^−154^ and 1.30, 1.29 to 1.32, P<1×10^−300^, respectively). In multivariable MR after accounting for adult body size, there was strong evidence of an effect between higher genetically predicted childhood body size and reduced fat mass index (beta, 95% CI: -0.05, -0.08 to -0.02, P=5×10^−4^). There remained strong evidence of an effect between higher genetically predicted adult body size and increase in fat mass index, after accounting for childhood body size (beta, 95% CI: 1.33, 1.30 to 1.35, P=1×10^−300^). Further estimates, including those stratified by sex, are in S5 Table.

The relationship between childhood body size and the odds of fracture in later life increased marginally, after accounting for lean mass index (OR, 95% CI: 0.82, 0.73 to 0.92, P=0.001) and reduced after accounting for fat mass index (OR, 95% CI: 0.76, 0.68 to 0.84, P=0.008) (S6 Table). This suggests that the childhood body size measure used in this analysis does not discriminate between adiposity and lean mass.

#### Sex-differential participation bias

There was evidence of an association between childhood and adult body size on sex. For example, results indicate that childhood and adult body size reduced the odds of being female (OR, 95% CI: 0.94, 0.89 to 0.99, P=0.029 and OR, 95% CI: 0.96, 0.91 to 1.00, P=0.049, respectively) (S7 Table). These artifactual relationships observed suggest some sex-differential participation bias is apparent.

#### Family-based analyses

Between-family univariable and multivariable estimates reveal consistent directions of effect with estimates from the whole population. Results are underpowered, however. This is due to i) the large reduction in the sample size, from the whole population (n=501,550) to a subset of siblings in the UK Biobank (n=39,507) and ii) there being difference in genetics in between-sibling analyses (S8 Table).

## Discussion

In this MR study, we employed a lifecourse framework to evaluate the effects of genetically proxied childhood and adult body size on fractures in later life. We observed strong evidence of an effect between higher childhood body size and reduced fracture risk in later life. This association became stronger after accounting for adult body size. Conversely, we identified a strong causal effect of adult body size on an increase in the odds of fracture in univariable as well as multivariable analyses, after accounting for childhood body size. Findings from this study suggest that higher body size in childhood may have a lasting influence on fracture risk in later life and therefore, the protective effect estimates observed in previous clinical and observational epidemiological research between BMI in adults and fracture risk (12, 13), are likely attributed to childhood effects. Where greater body size leads to an adaptive change in bone size and strength during growth, our results suggest that this does not occur in later life once growth has ceased. Importantly, the childhood body size measure used in this investigation does not discriminate between adipose and lean mass, or between fat stored in different compartments of the body. Furthermore, investigating birthweight as a third exposure showed that the effects of childhood and adult body size observed were not a result of body size at birth.

We additionally investigated whether plausible risk factors for fractures served as intermediate variables (mediators) on the causal pathway between childhood body size and fracture risk in later life, using two-step MR. Genetically predicted childhood body size was strongly associated with a decrease in calcium and vitamin D levels and an increase in eBMD in adulthood. There was, however, very little evidence of an effect between vitamin D and calcium on the odds of fracture in later life. On the other hand, we observed a strong causal association between eBMD and a decrease in the odds of fracture risk. Indeed, adiposity in childhood has been shown previously to be causally related to BMD in childhood, specifically of the limbs, pelvis, and spine, and not the skull (16). One hypothesis is that this reflects the positive effects of loading on bone formation at weighted sites. The effect we show between childhood body size and eBMD suggests that body size affects the amount of trabecular bone. As well as there being a scaling relationship between body size and overall bone size during growth, the internal bone structure is positively influenced by body size. This appears to persist throughout life, protecting against fracture risk regardless of BMI reduction in adulthood. In addition, whilst calcium and vitamin D supplementation is recommended for fracture prevention (49-51), findings from randomised clinical trials yield conflicting conclusions regarding their efficacy (52-54). Our results are supported by a recent MR study showing that genetic predisposition to lower levels of vitamin D and estimated calcium intake from dairy sources did not appear to be associated with fracture risk (19). Furthermore, MR investigations have shown that genetic evidence of increased serum calcium levels did not improve eBMD (55, 56).

Since effects of body weight are often sex-specific (27), we investigated the relationship between childhood body size and fracture risk as well as childhood body size and eBMD, separately by sex. The strength of the genetically predicted effect of childhood body size in males compared to females was more than 2-fold in magnitude on both outcome measures. Upon accounting for adult body size, these differences diminished, suggesting that our adult body size measure was, to a large degree, responsible for the sex differences observed in the outcomes of interest. Furthermore, higher childhood body size was strongly associated with a decrease of bioavailable testosterone in males and increase in females. This finding is supported by the literature, whereby obese males have been characterised by a decrease in testosterone levels with increasing body weight (57). Their female counterparts, conversely, have been shown to develop a condition of functional hyperandrogenism (58), which is, in most cases, detectable by testosterone elevation (57). Reduced SHBG synthesis and circulating blood levels have also been shown to represent the sole common mechanism response for this in both males and females (57), with the former observed in our findings as additionally occurring in response to higher body size in childhood. There was also strong evidence that bioavailable testosterone reduced the risk of fractures in both males and females. These associations, again, were stronger in males than they were in females. Furthermore, since the sex hormone measures used in this study were quantified in an adult population (mean age: 56.5 years), it is likely their effect is more strongly related to adult body size. This is in line with the literature, which has shown age-related testosterone deficiency to be the most important factor of bone loss in elderly men (59) and that SHBG in midlife is linked with injury risk in both sexes (28). It is additionally plausible, that the sex-differential associations observed in this study are, in part, a result of sex-differential participation bias, where the determinants of study participation affect females and males to differing extents (46). Evidence of this has been shown where artifactual associations between variants associated with childhood and adult body size and sex were observed.

From a public health perspective, the relationship between BMI and fracture risk is complex, as obesity remains a major risk factor for co-morbidities, including diabetes, cardiovascular diseases, cancer, and other health problems that may lead to further morbidity and mortality (60). It is nevertheless of importance to quantify the effect of BMI at different stages in the lifecourse on the risk of fracture in later life, to help i) identify modifiable pathways to fracture risk to decipher potential intervention targets, and ii) enhance the predictive value of BMI at different time points in fracture risk case finding (13, 60). In addition, this investigation highlights the importance of processes operating across the lifecourse, that influence the development of risk in later life (61).

### Strengths and limitations

Investigations have previously assessed the relationship between BMI and fracture risk observationally, as well as cross-sectionally, using MR methods. This is a unique study in that it estimates the effects of body size on fracture risk at separate timepoints in the lifecourse, whilst inferring causality by utilising the relationship between genetic variants robustly associated with a modifiable exposure or biological intermediate of interest and a disease outcome. An important and perhaps underreported methodological limitation in much of the obesity literature is through the use of BMI as an imperfect measure of adiposity (23). Whilst BMI indicates overweight relative to height, it does not discriminate between adiposity and lean mass. This study explores this measure to conclude that it is both indicative of adiposity and lean mass. This study additionally revealed potential mediators on the causal pathway between childhood body size and fracture risk, to aid in the identification of prospective intervention targets that may help to reduce fracture risk in later life. Furthermore, this investigation was able to leverage large sample sizes available through the UK Biobank study (n=453,169) for all measures used, by calculating expected bias and Type 1 error rate under the null that could result from using overlapping samples. In addition, weighted median and MR-Egger methods were used to assess the robustness of univariable results to horizontal pleiotropy.

This study, however, also has important limitations. First, self-reporting of body size in childhood by participants may have led to differential social desirability bias, in relation to retrospective weight recall at age 10. Moreover, the age of participants in adulthood when reporting this information could have influenced this measurement. To account for this, GWAS were computed on individuals who had both measures available adjusting for age, as well as sex and the genotyping chip. Second, our measure of childhood body size did not discriminate between adipose and lean mass, nor did it between fat stored in different compartments of the body. Third, using sex hormones quantified in adulthood, as opposed to childhood, where sex hormone levels are substantially different, limited our ability to decipher potentially important mechanisms between childhood body size and sex hormone regulation at the same timepoint. We additionally used fat and lean mass measures form adulthood, which likely associated more strongly with adult BMI than childhood as a result. Fourth, in using the UK Biobank, selection bias is a central limitation. Participation in the UK Biobank has been shown to be associated with being older, female and living in areas that are less likely to be socioeconomically deprived than individuals in nationally representative data sources (62). Therefore, this analysis is under-representative of younger, male, non-binary or any other gender identity individuals as well as those from the lowest socioeconomic position group. This has the potential to result in problems for instrumental variable analyses (63). In addition, we have shown that our sex-stratified results may, in part, exhibit artefactual autosomal heritability in the presence of sex-differential participation bias, which has been shown to lead to incorrect inferences in downstream analyses (46). Moreover, since allele frequencies as well as risk factors and diseases vary between subgroups in the population, confounding is plausible. This study thus performs analyses in homogeneous populations of European ancestry (64), therefore only depicting effects within this single ancestry group that may not be generalisable to other ancestry populations. Future research would benefit from replicating this across a broader range of different ancestries.

## Conclusions

This investigation provides novel evidence that genetically proxied childhood body size has a direct effect on reduced fractures in later life, via increased eBMD. Importantly, findings additionally indicate that higher BMI in adulthood is indeed a risk factor for fractures, opposing earlier clinical and epidemiological observational research findings which denote it as protective. The protective effect estimates previously observed between BMI in adults and fracture risk are therefore likely attributed to childhood effects.

## Data Availability

The primary data from the UK Biobank resource that support the findings of this study are accessible upon application (https://www.ukbiobank.ac.uk/).

## Acknowledgements

We would like to thank the UK Biobank study and all participants who contributed to it, as well as the authors of all the GWAS who made their summary statistics available for the benefit of this work. Estimates on childhood and adult body size were derived previously using data from the UK Biobank (app #15825).

## Financial Disclosure Statement

This work was in part supported by the Integrative Epidemiology Unit which receives funding from the UK Medical Research Council and the University of Bristol (MC_UU_00011/1). GDS conducts research at the NIHR Biomedical Research Centre at the University Hospitals Bristol NHS Foundation Trust and the University of Bristol. The views expressed in this publication are those of the author(s) and not necessarily those of the NHS, the National Institute for Health Research or the Department of Health. GMP is supported by the GW4 Biomed Doctoral Training Programme, awarded to the Universities of Bath, Bristol, Cardiff and Exeter from the Medical Research Council (MRC)/UKRI (MR/N0137941/1). TGR was a UKRI Innovation Research Fellow whilst contributing to this study (MR/S003886/1). TMF has received funding from the Medical Research Council (MR/T002239/1) EU-IMI SOPHIA and GSK. JT is supported by an Academy of Medical Sciences (AMS) Springboard award, which is supported by the AMS, the Wellcome Trust, GCRF, the Government Department of Business, Energy and Industrial strategy, the British Heart Foundation and Diabetes UK (SBF004\1079). The funders had no role in study design, data collection and analysis, decision to publish, or preparation of the manuscript.

## Notes

### Competing Interest Statement

Tom G Richardson is employed part-time by Novo Nordisk outside of this work. All other authors declare no competing interests.

### Author Declarations

The UK Biobank resource (https://www.ukbiobank.ac.uk/).

